# Can You Beat the Music? Validation of a Gamified Rhythmic Training in Children with ADHD

**DOI:** 10.1101/2024.03.19.24304539

**Authors:** Kevin Jamey, Hugo Laflamme, Nick E. V. Foster, Simon Rigoulot, Sonja A. Kotz, Simone Dalla Bella

**Author notes:** Correspondence concerning this article should be addressed to Kevin Jamey  and Simone Dalla Bella. Kevin Jamey and Hugo Laflamme are co-equal first authors contributors.

## Abstract

Neurodevelopmental disorders like ADHD can affect rhythm perception and production, impacting the performance in attention and sensorimotor tasks. Improving rhythmic abilities through targeted training might compensate for these cognitive functions. We introduce a novel protocol for training rhythmic skills via a tablet-based serious game called Rhythm Workers (RW). This proof-of-concept study tested the feasibility of using RW in children with ADHD. We administered an at-home longitudinal protocol across Canada. A total of 27 children (7-13 years) were randomly assigned to either a finger-tapping rhythmic game (RW) or a control game with comparable auditory-motor demands but without beat-synchronization (active control condition). Participants played the game for 300 minutes over two weeks. We collected data (self-reported and logged onto the device) on game compliance and acceptance. Further, we measured rhythmic abilities using the Battery for the Assessment of Auditory Sensorimotor and Timing Abilities (BAASTA). The current findings show that both games were equally played in duration, rated similarly for overall enjoyment, and relied on similar motor activity (finger taps). The children who played RW showed improved general rhythmic abilities compared to controls; these improvements were also positively related to the playing duration. We also present preliminary evidence that executive functioning improved in those who played RW but not controls. These findings indicate that both games are well-matched. RW demonstrates efficacy in enhancing sensorimotor skills in children with ADHD, potentially benefiting executive functioning. A future RCT with extended training and sample size could further validate these skill transfer effects.

---

Engaging in music learning proves to be a rewarding and captivating pursuit (Mas-Herrero et al., 2021; Vuust et al., 2022; Zatorre, 2015) for typically developing children but also for children following atypical developmental trajectories (Fasano et al., 2022; Kim et al., 2009; Kraus et al., 2014; Wilde & Welch, 2022). More generally, intrinsically pleasant activities, such as learning music, yield greater learning outcomes and psychological well-being than externally driven tasks motivated by external incentives, such as receiving praise or good grades (di Domenico & Ryan, 2017; Ryan and Deci, 2017). Music is widely recognized as an effective rehabilitative tool for many neurodevelopmental and acquired or progressive neurological conditions, including autism (Latif et al., 2021; Sharda et al., 2018, 2019), attention-deficit/hyperactivity disorder (ADHD; Zhang et al., 2017), dyslexia (Flaugnacco et al., 2015), and Parkinson’s disease (e.g., Puyjarinet et al., 2022; Dalla Bella et al., 2017; Hackney & Earhart, 2009).

Training musical abilities is a multifaceted process engaging processes like auditory-motor skills and cognitive processes (e.g., attention and memory). In particular, auditory-motor integration, the interplay of auditory perception and action, holds a pivotal role in monitoring attention and behavior, influencing focused attention to dynamic signals such as audition and speech when navigating complex environments (Alho et al., 2014; Liu et al., 2020; Wilkinson et al., 2020). Learning a musical instrument engages these processes, integrating physical actions with auditory cues to produce music. Auditory-motor skills are fundamental in early child development (Trainor & Cirelli, 2015) and are integral for mastering a musical instrument in adulthood (Lega et al., 2016; Wollman et al., 2018; Zatorre et al., 2007). The systematic practice of auditory-motor functioning is essential during the learning and practicing phases of a musical instrument, contributing to the acquisition or restoration of mental and physical skills (Grau-Sánchez et al., 2022). Music training is known to establish robust brain connections between the auditory and motor systems (Dalla Bella et al., 2024; Herholz & Zatorre, 2012; Merrett et al., 2013; Strait et al., 2014; Wan & Schlaug, 2010). These connections, in turn, expand to the sensorimotor networks (Karpati et al., 2016; Li et al., 2018), executive functioning (Degé & Frischen, 2022), speech and language (Gordon et al., 2015), and socio-emotional processing (Gaudette-Leblanc et al., 2021).

## Rhythmic training in auditory-motor integration

In a musical context, auditory-motor integration often corresponds to producing motor actions in response to highly predictable sequences, particularly when we synchronize movement to the pulse of the music (i.e., its *beat*). A musical pulse is the regular and recurring underlying time frame in a musical piece, often expressed by the “tempo” in beats per minute. The ability to accurately perceive and time motor actions to the temporal regularity of a stimulus is widespread in humans, including children, and is linked with the general idea of temporal prediction (Sowiński & Dalla Bella, 2013; Tranchant et al., 2016; Honing, 2012; Grondin, 2010; Kotz et al., 2018). We continuously predict future events based on information retrieved from patterns and repetitions we perceive in the external world (Büchel et al., 1999; Coull et al., 2011; Rao & Ballard, 1999; Schwartze et al., 2013). Temporal predictions are linked to self-sustained internal oscillations that align with the rhythmic features of an incoming stimulus sequence (Fujioka et al., 2012; Large & Jones, 1999; Nozaradan, 2014). This fundamental process, known as “entrainment”, creates time-based predictions that impact motor coordination where movements are adjusted to match the expected beat intervals. Aligning bodily motions with an evenly timed auditory beat is expected to enhance the “entrainment” process and likely affects the efficiency of temporal predictions recruited in other systems (e.g., higher-order language and attention learning; Rimmele et al., 2018). The refinement of prediction accuracy throughout the timing systems (e.g., Schwartze & Kotz, 2013) likely influences sensorimotor, attentional, and language functions that rely on efficient timing functions (Balasubramaniam et al., 2021; Dalla Bella, 2020; Patel & Iversen, 2014).

## Benefits of rhythmic training

Synchronizing perception and action to a beat is apparent in many motor activities, including finger or foot tapping, walking, or dancing (Dalla Bella et al., 2017; Fujii & Schlaug, 2013; Repp & Su, 2013; Sowiński & Dalla Bella, 2013; Tranchant et al., 2016). It involves neural networks tied to motor control (premotor area, supplemental motor area, basal ganglia, and cerebellum) and executive functions (Chen et al., 2008; Coull et al., 2011; Fujii & Wan, 2014; Grahn & Brett, 2007; Grahn & Rowe, 2009). In rhythmic synchronization tasks, maintaining consistent alignment with the musical beat requires executive functioning, such as inhibition control to resist disruptions from off-beat temporal information and set-shifting when switching between distinct sections of a musical piece (Jamey et al., 2023; Vuust et al., 2011; Zuk et al., 2014). Consequently, rhythmic training may improve these specific executive functions, and recent findings in pre-schoolers show a greater impact of rhythmic training on inhibition control than pitch-based training (Frischen et al., 2019). Overall, rhythmic training is a promising and engaging new entry point for gaining and regaining motor and attentional functioning.

## Gamified rhythmic training

Many gamified solutions offer effective rehabilitation by providing purpose-driven programs accessible on mobile devices for at-home treatment and remote progress tracking (Agres et al., 2021). In particular, “serious games” are interactive digital applications designed to provide an engaging and entertaining experience while simultaneously serving educational, training, or informational objectives. These games enhance behavioral research inclusivity, rigor, and reproducibility (Long et al., 2023). Recent reviews and meta-analyses confirm the success of “serious games” in rehabilitating cognitively impaired adults and addressing neurological conditions like dementia, Alzheimer’s, and schizophrenia (Abd-alrazaq et al., 2023; Francillette et al., 2021; Saragih et al., 2022). Extensive research, outlined in a review of 145 studies by Kokol et al. (2019), highlights the promise of “serious games” focusing on beat perception and synchronization in children with developmental disorders. Rhythm-based games have shown promise in addressing deficits in sensorimotor and attention-related functions observed in conditions such as autism, ADHD, dyslexia, and stuttering, as well as Parkinson’s Disease (Bégel et al., 2022; Dauvergne et al., 2018; Falk et al., 2015; Puyjarinet et al., 2022; Srinivasan et al., 2015). Combining music and gamified approaches, which activate the dopaminergic system, creates a particularly motivating training environment (Koshimori et al., 2019). The effects of rhythmic training on the dopaminergic system are particularly relevant for conditions where this system is deregulated, such as Parkinson’s disease and ADHD. Impaired rhythmic and non-rhythmic mechanisms seem to overlap in several neurological disorders, pointing toward the beneficial effects of rhythmic training upon non-rhythmic processes (Bégel et al., 2017; Pasqualotto et al., 2021; Puyjarinet et al., 2017; Srinivasan et al., 2016).

## Rhythm Workers

Despite the large number of off-the-shelf rhythmic games on the market, there is a lack of gamified approaches to rhythmic training suitable for a serious game (see Begel et al., 2017, for a review). To fill this gap, we recently developed a game named “Rhythm Workers” (RW), specifically designed to train rhythmic skills (Bégel et al., 2018). We tailored the first prototype of the game to an adult population for training patients with a neurodegenerative disorder (Parkinson’s disease; Dauvergne et al., 2018; Puyjarinet et al., 2022). The game’s premise is that rhythmic abilities can improve via a training program using finger tapping to music, with beneficial effects for motor control and cognition (e.g., (Dalla Bella, 2022). In RW, participants play by tapping their fingers to the beat of rhythmic auditory stimuli (e.g., music) on a tablet. The objective is to construct a building under a specific time limit that varies for each game level. The construction progress, aesthetics, and player score depend on the accuracy and consistency of the alignment between the tapping times and the stimulus beat. The initial game prototype featured progressively challenging levels correlated with beat saliency of a chosen auditory stimuli (for specifics, refer to Bégel et al., 2018). The game’s first iteration underwent testing in healthy adults (Bégel et al., 2018) and in participants with Parkinson’s disease as part of an intervention protocol, comparing RW to an active control group (Puyjarinet et al., 2022). So far, the game has not been used or validated in neurodevelopmental populations.

## Main objectives

This study aimed to assess whether children with ADHD engage with and reliably adhere to a 2-week training protocol using a child adaptation of RW. The protocol was conducted remotely, including at-home training sessions, online testing, equipment mailing, and teleconferencing. The primary objective was to determine if this protocol enhances rhythmic abilities compared to an active non-rhythmic control game. Additionally, the study sought preliminary evidence for improved executive functioning after rhythmic training. ADHD, which impacts 2 to 7% of school-aged children, manifests core symptoms such as hyperactivity, impulsivity, and inattention (Erskine et al., 2013; Polanczyk et al., 2014; Willcutt et al., 2012), which are frequently associated with timing-related issues (Gustafsson et al., 2023). Evidence collected over the past two decades also shows that ADHD is associated with poor duration-based timing abilities and difficulties in tracking rhythmic information (Noreika et al., 2013; Puyjarinet et al., 2017). The concurrent rhythmic and attentional deficits observed in ADHD suggest that there may be a strict relation between timing and executive functioning impairments in ADHD, notably for inhibition control and cognitive flexibility, pointing to potentially shared mechanisms. Impulsiveness is a timing and motor-based impairment whereby response styles are premature, impatient, delay-aversive, and non-reflected (Puyjarinet et al., 2017; Rubia, 2002; Rubia et al., 2009). If these systems are causally linked, improving rhythmic functions through training, such as with RW, might benefit attention-based executive functions. This study serves as a first proof-of-concept to validate the use of a rhythmic “serious game” for children with ADHD, laying the groundwork for future randomized control trials exploring its effects on timing and executive functioning.

## Specific aims and hypotheses

### Aim 1

To examine the feasibility of using RW in children with ADHD, compared to an active control condition, namely a non-rhythmic serious game (Frozen Bubble). We focussed on compliance with the protocol and game acceptance (as defined in the Methods section) and expected them to be sufficient and comparable between training conditions.

### Aim 2

To investigate RW’s efficacy in enhancing rhythmic abilities compared to the control game. Here, we expected better rhythmic performance for those who played RW than the control game.

### Exploratory Aim

A corollary objective was to explore whether RW improved executive functioning, focusing on inhibition control and cognitive flexibility, which are central to sensorimotor synchronization training. We expected better executive functioning performance for those who played RW than the control game.

## Methods

### Participants

The study involved 27 children (aged 7-13 years, 3 females) with ADHD recruited through announcements, advertisements, and targeted social media outreach within the ADHD community. We included participants aged between 7 and 13 years and fluent in either French or English, with 60% of participants choosing to participate in French. All participants had an ADHD diagnosis based on a written declaration from a parent; however, we did not ask for an official clinical report. We also excluded participants if parents reported comorbid neuropsychological, psychiatric, or developmental disorders related to ADHD and more than two years of formal musical training. Overall, 96% of the parents provided information on their child’s video gaming history and socio-economic status. Weekly video game playing was assessed on a two-point Likert scale (0 = No; 1 = Yes), along with the weekly playing duration. Z-scores were calculated for these items and combined into a composite overall video-gaming history score. Socio-economic status was determined by converting household income and education level into Z-scores, which were then combined. Participants received $50 upon completing the protocol. Due to the small sample size in this proof-of-concept study, non-parametric statistical analyses (Wilcoxon rank sum tests) were employed for group comparisons related to the participants’ backgrounds. See Table 1 for demographic and background characteristics.

**Table 1.**
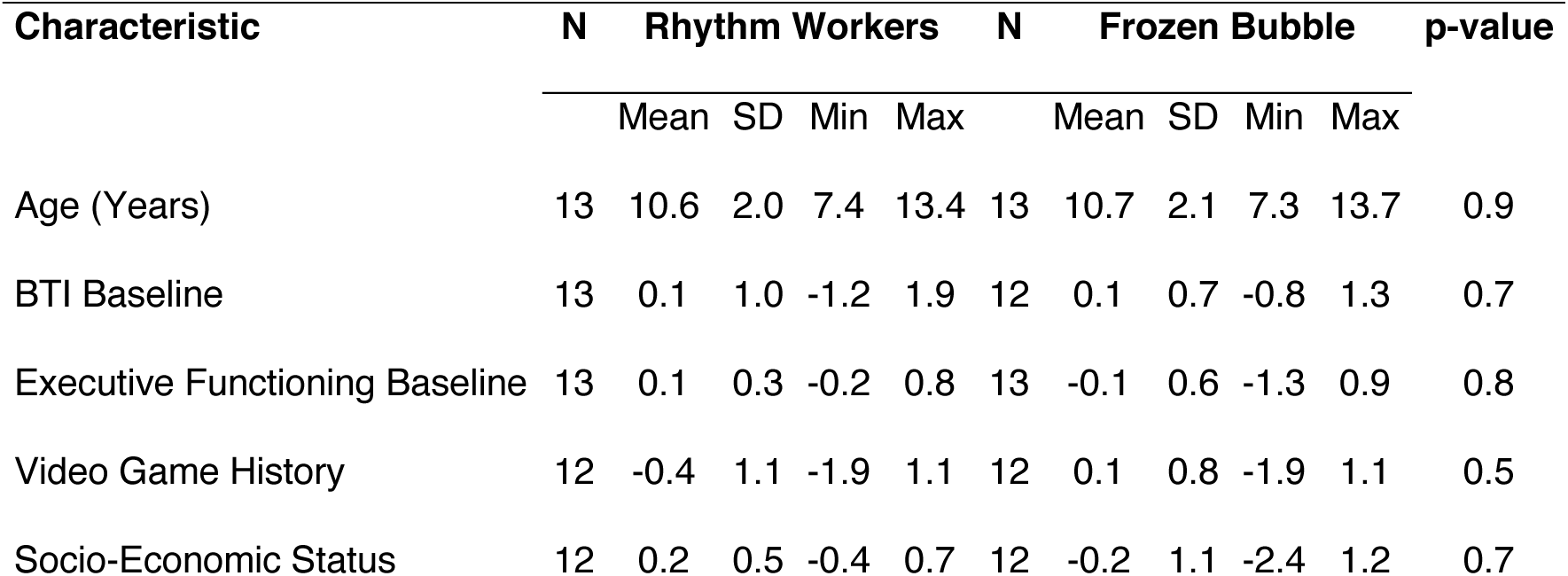
Participant Background and Performance Baselines.

| Characteristic | N | Rhythm Workers |  |  |  | N | Frozen Bubble |  |  |  | p-value |
| --- | --- | --- | --- | --- | --- | --- | --- | --- | --- | --- | --- |
|  |  | Mean | SD | Min | Max |  | Mean | SD | Min | Max |  |
| Age (Years) | 13 | 10.6 | 2.0 | 7.4 | 13.4 | 13 | 10.7 | 2.1 | 7.3 | 13.7 | 0.9 |
| BTI Baseline | 13 | 0.1 | 1.0 | -1.2 | 1.9 | 12 | 0.1 | 0.7 | -0.8 | 1.3 | 0.7 |
| Executive Functioning Baseline | 13 | 0.1 | 0.3 | -0.2 | 0.8 | 13 | -0.1 | 0.6 | -1.3 | 0.9 | 0.8 |
| Video Game History | 12 | -0.4 | 1.1 | -1.9 | 1.1 | 12 | 0.1 | 0.8 | -1.9 | 1.1 | 0.5 |
| Socio-Economic Status | 12 | 0.2 | 0.5 | -0.4 | 0.7 | 12 | -0.2 | 1.1 | -2.4 | 1.2 | 0.7 |

### General procedure

This single-blind, two-arm, parallel-group randomized control study assigned participants to the experimental group (playing the rhythmic game, RW) or the active control group (playing the non-rhythmic game, FB). The general procedure comprised a screening phase, remote testing and training, and a final debriefing interview with parents.

### Screening

A Zoom teleconferencing interview was conducted to ensure understanding and compliance with inclusion and exclusion criteria. Eligible participants were enrolled in the study once they consented via email.

### Remote testing and training

The experiment was conducted between October 2021 and March 2022 across Canada. The entire research protocol was conducted remotely from the participants’ homes, using the BRAMS Online Testing Platform, teleconferencing video meetings (Zoom), and testing equipment mailed to the participants. The equipment was shipped to the participants considered eligible for the study. Rhythmic and cognitive abilities of each participant were assessed before and after the 2-week training program via four video meetings. Upon reception of the equipment, two video meetings (separated by at least 90 minutes) were scheduled with the participant. During the first meeting, participants completed four online executive functioning tasks on the BRAMS Online Testing Platform (https://brams.org/online-testing-platform-training/), comprising Go/No-Go, Flanker, and Set-Shifting (see below for details), and an N-back working memory task (not included in the present analyses). During the second meeting, participants completed tablet-based rhythmic tests from the Battery for the Assessment of Auditory Sensorimotor and Timing Abilities (BAASTA; (Dalla Bella et al., 2024; Dalla Bella, Farrugia, et al., 2017); see below for details). Participants were instructed not to reveal which game they had been assigned to the experimenter and not to use the tablet before the rhythmic task session. The researchers conducting executive functioning and rhythmic tests were blinded to the participants’ group assignments. At the end of the second meeting, a research assistant (non-blinded) presented and explained to participants how to play the game they received. Participants were instructed to begin playing the game once they understood the instructions explained by the research assistant. The third and fourth video meetings, comparable to the first two meetings, were scheduled after the end of the training period. At the end of the fourth meeting, the research assistant scheduled a debriefing meeting to collect general comments and feedback about the game.

### Randomization

Participants were randomized using covariate adaptive randomization with the “minimization” approach (Lin et al., 2015) by an experimenter who was not involved in any other task during the protocol execution. The first 6 participants were assigned via block randomization (random permutation of 3 in the experimental group and 3 in the control group) without regard to covariates. The seventh and eighth participants received purely random assignments. The remaining participants were then assigned using “biased coin” randomization with p = .8 (Pocock & Simon, 1975) to minimize group imbalance in the number of participants, gender, age, language (English / French), and music experience. These variables were group-matched at p > .89 in the final sample.

### Equipment

A Samsung Galaxy Tab A 8.0” 2019 Android tablet (model SM-T290) was sent to participants for use in the experiment. The tablet had a 2GHz processor, 1280 x 800 pixels display resolution, and 32 GB of RAM. Participants used the touch screen to navigate and play the applications. JBL Tune 500 Wired On-Ear Headphones with One-Button Remote/Mic were used to play music and sounds from the applications. All materials required to complete the study were mailed using postal and courier services. This material included the tablet (the game assigned to the participant, and the rhythmic assessment application BAASTA), a gooseneck phone holder, headphones, paper questionnaires, game-specific instructions, and sanitizing kits.

### Training protocol

Participants were instructed to play their assigned game 30 minutes a day, five days a week, for two weeks (300 minutes of gameplay). They were allowed to combine two 30-minute sessions into a 1-hour session to provide more flexibility in planning. Participants received the following instructions: *“I would like you to play it for about 30 minutes a day, five days a week, for 2.5 hours a week. You can play more if you want, but don’t play more than an hour a day”.* No child reported playing more than one hour a day. During the two weeks of playing the game, participants were asked to complete a questionnaire before and after each session. The questionnaire included nine yes or no questions about their mood taken from a children’s mood questionnaire (Derbaix & Pecheux, 1999). Participants also indicated their progress in the game (level number), as well as the perceived difficulty of the game on a 5-item Likert scale (1 = very easy; 5 = very hard) and how much they enjoyed the game on a scale of (1 = very boring; 5 = very fun).

### Stimulus selection

Before the training, we conducted a pilot study in typically developing children to select the musical stimuli for the rhythmic game. Fifty-eight musical excerpts (30 seconds each) were chosen to be appealing to children between the ages of 7-13 years, with tempi ranging from 78 bpm to 135 bpm, and including an equal proportion of electronic, classical, pop-rock, jazz-funk, and traditional genres. All musical excerpts were instrumental and did not include vocals. Half of the excepts were composed by musicians for the commercial release of RW (BeatWorkers, commissioned by BeatHealth Company). The other half of the stimuli were selected from a royalty-free music archive composed by Kevin MacLeod (https://incompetech.com/music/royalty-free/faq.html). Two co-authors (XX and YY) are professional musicians and disc jockeys who rated each stimulus for the percentage of syncopation. Syncopation was calculated by calculating the percentage of the beat with a low or inexistant pulse clarity (little to no sound on the beat) over 16 beat increments. In an online experiment run on the BRAMS Online Testing Platform, we asked 25 typically developing children aged 7-13 years (mean age = 10.23, SD = 2.11; 54% females) to tap to the beat of each stimulus, using the space bar of a keyboard or by tapping on the green button on a touch screen.

At the end of each excerpt, children rated the stimuli on a 5-point Likert scale in terms of enjoyment (1 = very boring; 5 = very fun; M = 2.72; SD = 1.11; Range = 2.3-3-3) and tapping difficulty (1 = very easy; 5 = very hard; M = 2.89; SD = 0.96; Range = 2.5-3.23). We computed the synchronization consistency of tapping performance using circular statistics (i.e., vector length; see Sowinski and Dalla Bella, 2013; Dalla Bella et al., 2017). Synchronization consistency was measured using vector length and was logit-transformed to reduce data skewness, a common practice for synchronization data (e.g., Kirschner & Tomasello, 2009; Sowiński & Dalla Bella, 2013). Scores and ratings of each of the fifty-eight musical stimuli are presented in Table 1.

To select songs for the 32 levels of RW, we primarily relied on the synchronization consistency rankings shown in Table 1. Music by Kevin MacLeod was prioritized because synchronization consistency varied within a broader range (0.07 – 1.51) than the commercial music of BeatHealth company (0.45 – 1.54), and thus provided greater margin to manipulate rhythmic difficulty. Moreover, royalty-free licenses enhance accessibility, transparency, and openness for future research. Music from the BeatHealth company was selected in specific situations to avoid songs that received low ratings for stimulus appreciation or extremely high ratings of perceived difficulty, as well as to diversify tempo and genre during level progression. In total, 25 musical stimuli by Kevin MacLeod and 7 by the BeatHealth company were used. The final order of the 32 selected songs across game levels was adjusted by XX based on their professional judgment.

**Table 2.**
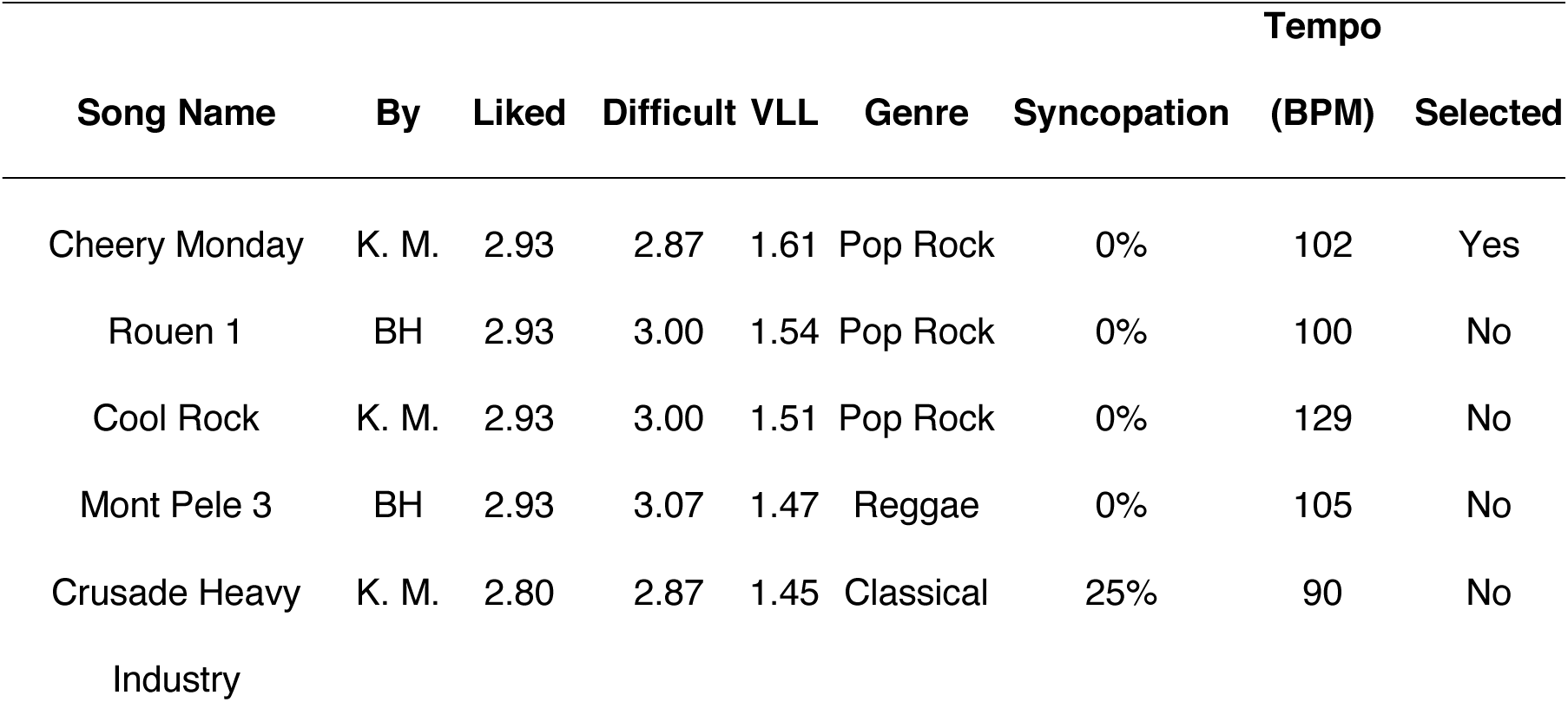
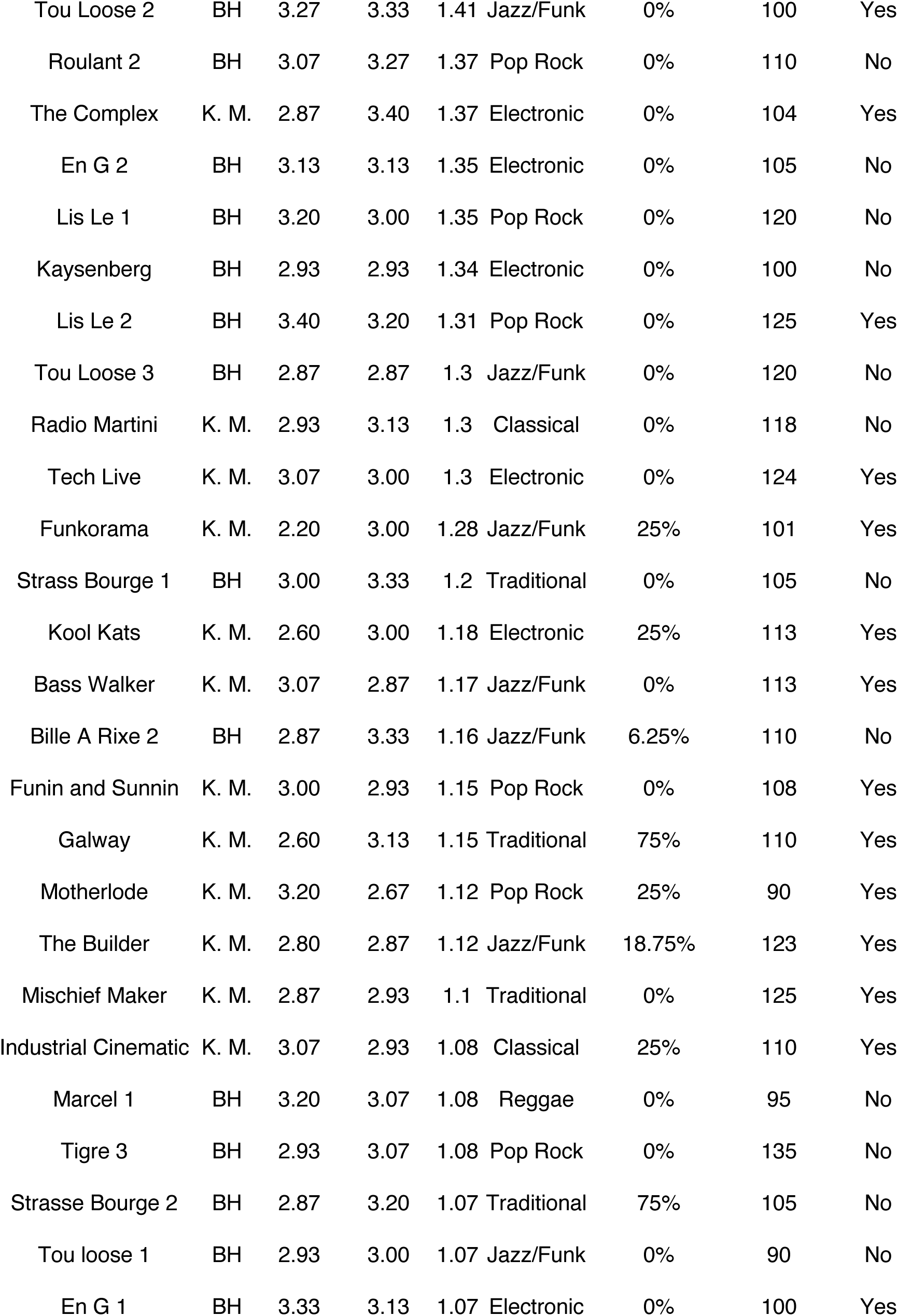
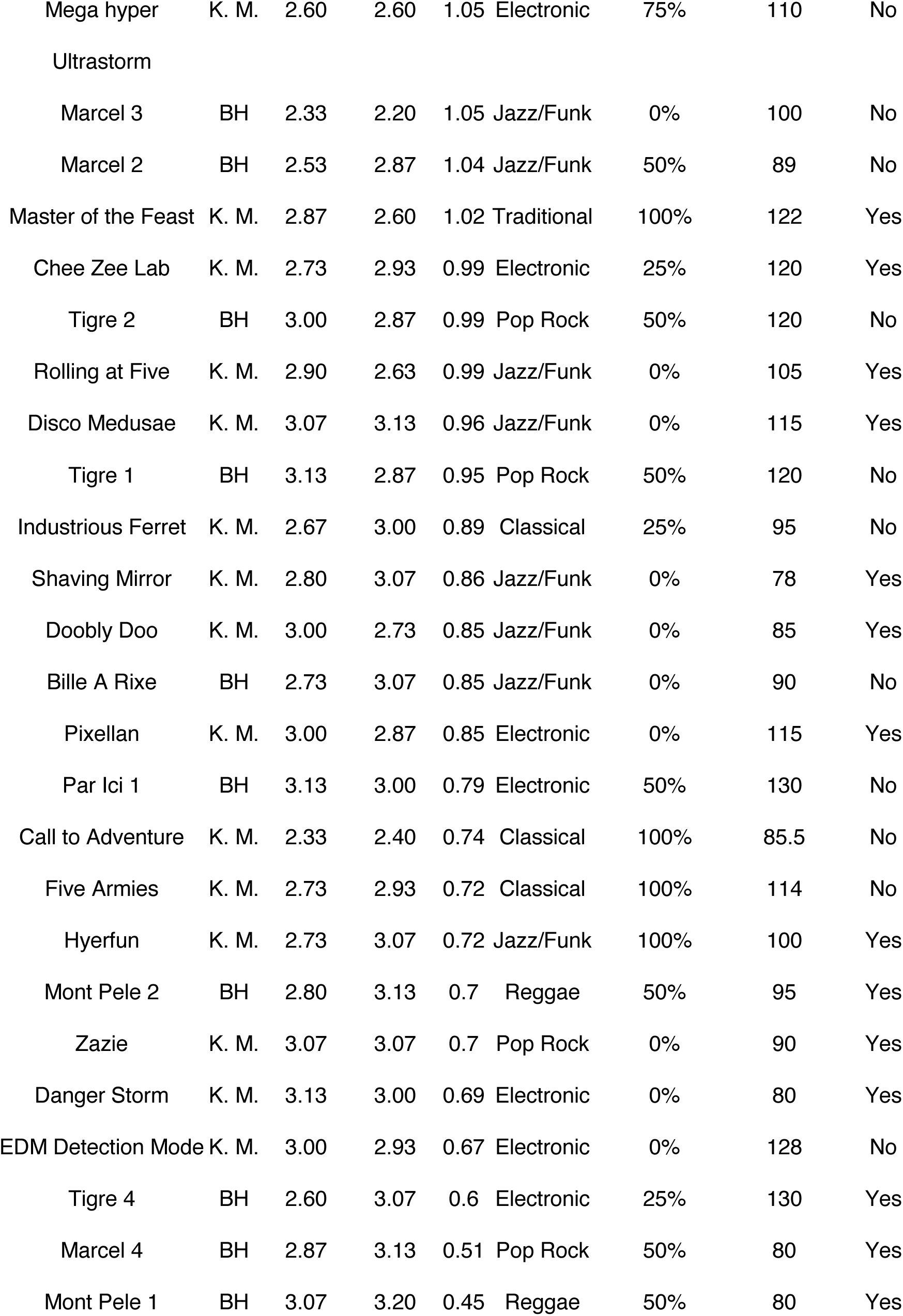
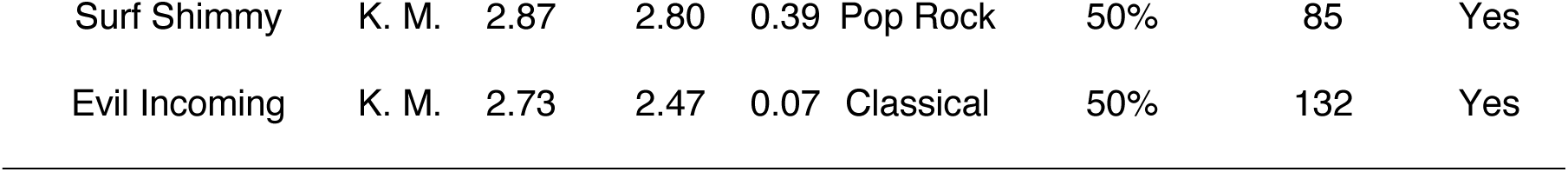
Description of musical excerpts, synchronization consistency (logit-transformed vector length), genre, syncopation, tempo, and participants’ ratings of pleasantness and perceived difficulty (provided on a 5-point scale) are also reported.

### Rhythmic “serious game”

Rhythm Workers (RW), the “serious game” used in this proof-of-concept study was initially developed for adults (Begel et al., 2018). The scoring method and synchronization condition of the game were similar, but significant improvements were made in terms of the visual interface, the gameplay, difficulty, and musical stimuli to suit children. First, we changed the musical stimuli from simple midi-generated music (Begel et al., 2018) to complex and layered instrumentations performed, recorded, and designed by professional musicians. The objective of this modification was to make the musical stimuli more engaging for children. Second, we added distractors that fit the motor level and cognitive functioning of 7–13-year-old children. Third, the game’s scoring system was also altered so fewer points were required to build a complete building within each level’s timeframe. Finally, the graphics and interface were also improved, giving the game a finished feel like an online commercial game (for a version of the game for the public, see https://www.beatworkers.com).

In this version of RW, players were asked to synchronize their finger taps to the beat of musical excerpts. The goal of the game was to construct a building by aligning the taps to the musical beat. Circular statistics were employed using data from the most recent eight taps to evaluate real-time tapping performance. To derive a comprehensive score, the assessment measured synchronization consistency (expressed as vector length) and accuracy (expressed as a vector angle representing the extent to which tapping tended to lead or follow the stimulus beat). The scoring process involved multiplying synchronization consistency (ranging from 0 to 1) by 100, yielding a score within the 0 to 100 range. Note that achieving maximum consistency (1.0) is unattainable in human performance. Three points were automatically added, resulting in a perfect score of 100 for outstanding players. Scores were then adjusted based on synchronization accuracy. For vector angles exceeding 60 degrees, deductions were made every 10 degrees, with decreasing accuracy. For instance, between 60 and 70 degrees, 5 points were deducted from the final score, and between 70 and 80 degrees, 10 points were subtracted, following a similar pattern. Players sometimes tapped in antiphase (i.e., midway between the beats), which was considered an erroneous performance. Antiphase tapping was identified during the level assessment when the player’s synchronization accuracy deviated by at least 120 degrees from the beat, given that the antiphase corresponds to 180 degrees.

Players received feedback while playing at the end of each 8-bar sequence. If the score was above 90, the expression “ACE!” showed on the screen (see Figure 1); the building story was constructed faster, and players scored the most points. For a score between 70 and 90, players saw “BIG!”; the building was constructed normally, and players scored points. Scores below 70 showed the words “ Err!!”; no progress was achieved on building the stories, and no additional points were given. Players could receive medals at the end of each level based on their total points, and this unlocked more game levels to play. Successive levels each required a greater number of accumulated medals to be unlocked. Accordingly, during their progression through the game, players usually had a range of 2-4 new levels ahead that they could access.

**Figure 1.**
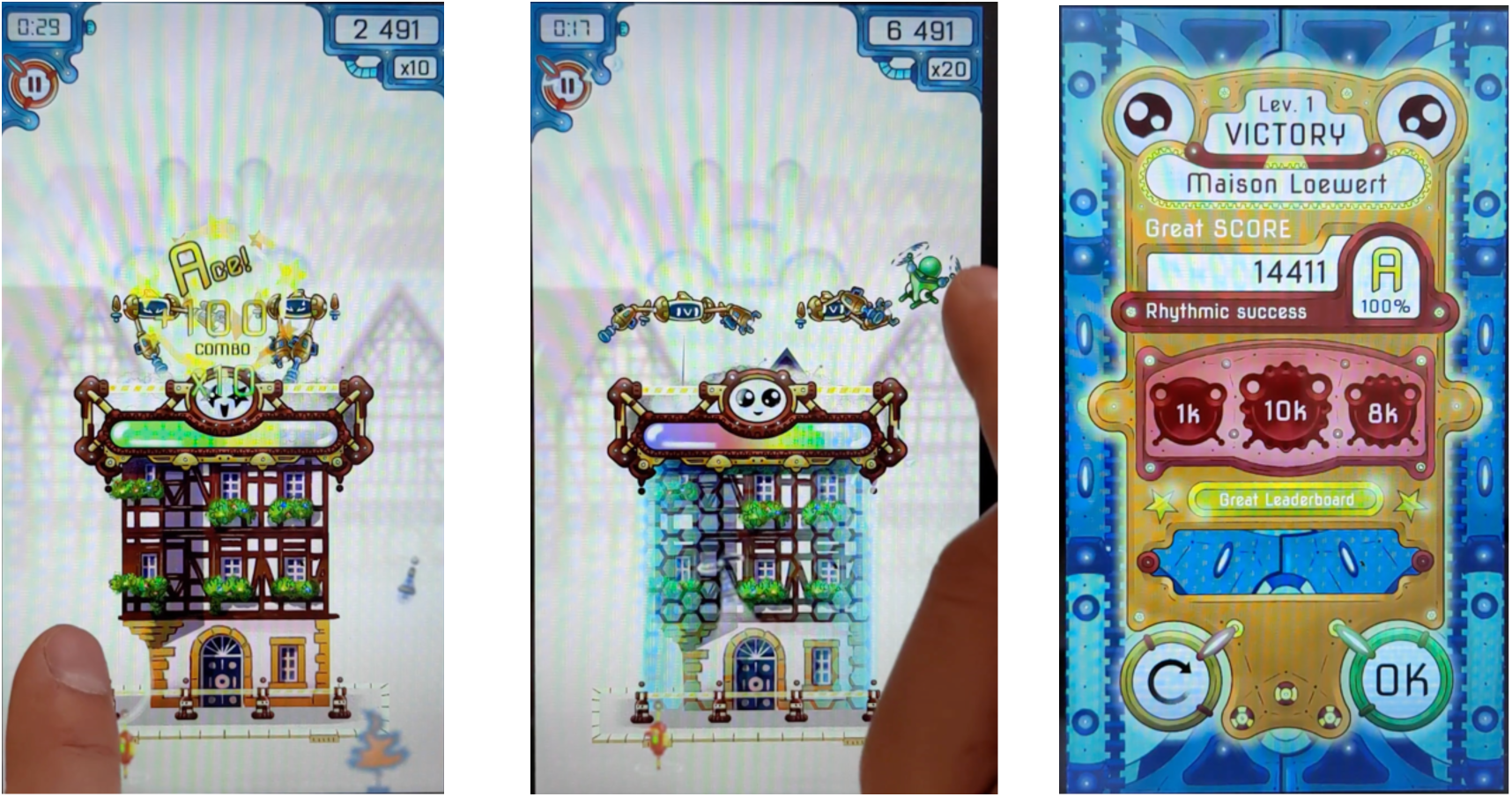
Example of Rhythm Workers gameplay (experimental training condition) used in this proof-of-concept study. On the left, a building was constructed, and the player scored maximum points and received visual feedback. In the center, the player tapped on a distractor on the beat. On the right, the player saw their scoreboard once the level was completed.

Prior to the start of this proof-of-concept study, the child-friendly, adapted version of RW was tested in another in-person laboratory survey in typically developing children between 6 and 13 years of age to ensure that the music selection, gameplay interface, and graphics were suitable for that age range. Feedback from 13 participants allowed us to improve the game and adaptation for children further. RW included 32 levels, each corresponding to one of the songs selected in the stimulus pilot study. The game’s progression was designed by increasing rhythmic difficulty using the synchronization consistency scores (vector length logit transform) obtained during the validation of the musical stimuli. Each level involved two repetitions of the same excerpt. The first repetition, a standard rhythmic task in which the participant taps to the beat of music without distractors, served as practice. The second repetition added moving “enemy” distractors that could be eliminated by tapping them on the beat. We varied distractor difficulty by increasing the speed (3 different speeds) at which they moved on the screen and how many taps were required to eliminate the enemy (1 or 2 taps). If the player did not destroy the distractor, the distractor attacked the building, reducing the player’s score. Distractor difficulty was introduced progressively in the game, and distractors appeared only in a sequence. During the first six levels, 1 to 2 “enemy” distractors appear, requiring 1 or 2 taps on the beat to eliminate them. The first six levels were limited to a maximum of 2 distractors so participants could familiarize themselves with the distractors. For the following levels, between 4 and 8 distractors appeared, requiring 1 or 2 taps on the beat to make them disappear. The number of “enemy” distractors was added for variety and to guarantee sustained attention throughout the training sessions. For this reason, we did not increase the number of distractors with increasing level progression but instead attributed the 4-8 distractors randomly per level. A sample of the musical stimuli of the game and examples of the gameplay can be found at the following link: https://osf.io/gykjd/?view_only=26be79399f1e48fe838107df437e0ceb

### Non-rhythmic serious game (active control game)

The control game, Frozen Bubble (FB) was based on a publicly available version of the game on GitHub (https://github.com/videogameboy76/frozenbubbleandroid) and is a puzzle arcade shooting game in which the goal is to clear the screen of descending bubbles using finger tapping. The arrangement of bubbles progressively moved down the screen, and the player lost if they reached the bottom of the screen. To make the bubbles fall, players had to arrange them by color. When three or more bubbles of the same color were stuck together, they fell off the screen, and any bubbles attached below became free (see Figure 2).

**Figure 2.**
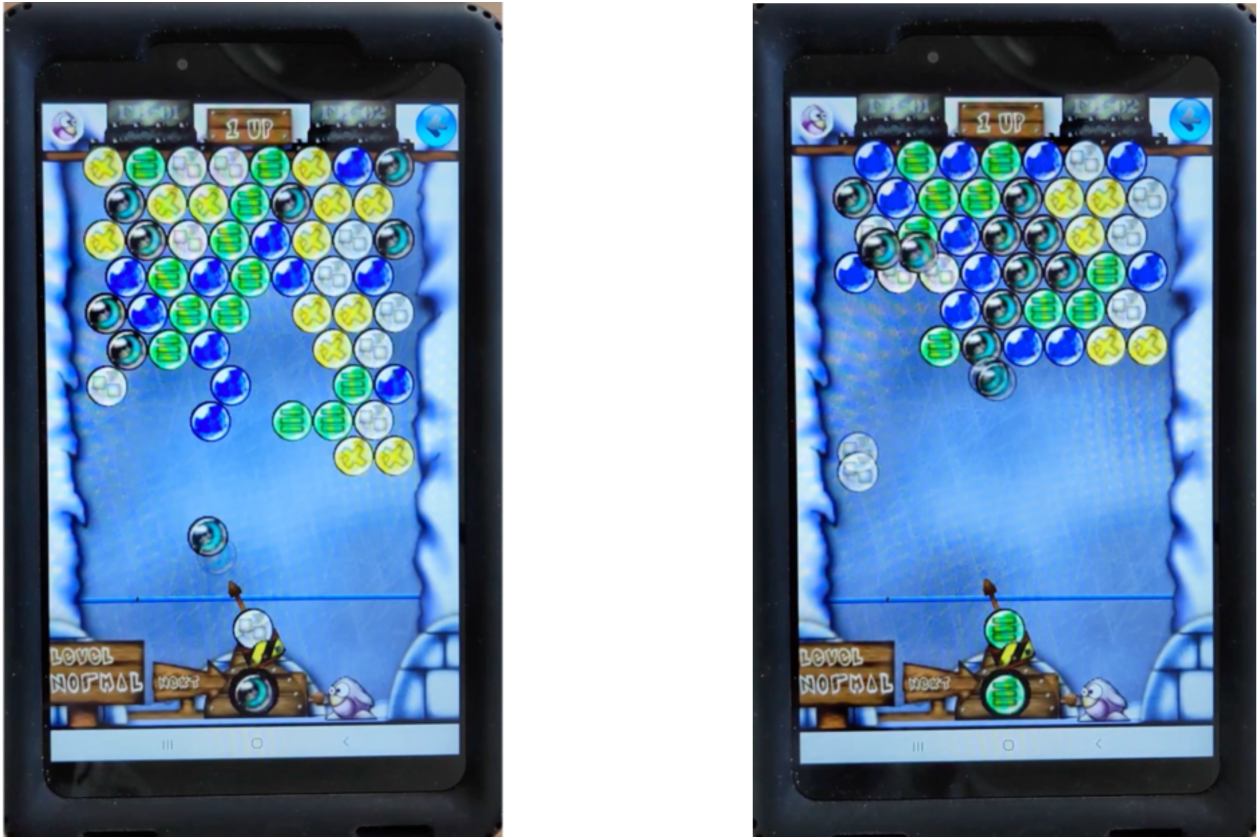
Example of Frozen Bubble gameplay (control training condition). On the left, the black bubble was released in the direction of the arrow by two finger taps (the first to arm, and the second to aim toward the higher two black bubbles and shoot). On the right, at a different moment in the game, a bubble has hit its target and freed bubbles.

The game played background instrumental music with a beat similar to RW, except players did not synchronize their motor movements to the beat. To make motor demands comparable with RW, we modified the FB game to double the number of taps (one to arm and one to launch the bubble gun), as described below. Players had a choice of three play modes: “Puzzle Mode”, “Arcade Mode, and “Player vs. Computer”. In “Puzzle Mode” an arrangement of bubbles of different colors was presented on the screen for each of the 100 levels. To launch a bubble, players had to tap twice on the place where they wanted to send the bubble: Once to load the gun and a second time to launch the bubble. This principle of shooting and connecting bubbles to clear them from the screen was the same across the three modes. In “Arcade Mode”, the bubble arrangement descended gradually on the screen, with new rows emerging from the top edge, and the player’s goal was to clear bubbles from the screen to survive as long as possible. In this mode, levels were not presented. In “Player vs. Computer” mode, an arrangement of colored bubbles was presented on the screen, and players played against the computer, which also had a screen with bubbles to eliminate; cleared bubbles were sent from the player to computer and vice-versa, adding to the counterpart’s arrangement of bubbles to be cleared. The game’s goal was to clear all bubbles from the screen before the computer cleared all bubbles from its screen. This was the most challenging mode.

### Evaluation of game compliance and acceptance

The first aim of this proof-of-concept study was to evaluate if participants adequately followed the study protocol and played the experimental and the control games in comparable ways.

### Compliance

The compliance levels in this study refer to the degree to which participants fulfilled the requested protocol targets. Complete compliance entailed using the assigned game for the requested time while respecting a two-week playing time distribution. We assessed participant compliance with protocol targets by comparing the cumulative training duration (i.e., of training sessions; self-reported and logged on the device) and CPT in minutes (i.e., time spent in active gameplay in game levels; logged on the device). Participant self-reports of session times and dates were cross-checked manually with logfiles for overall coherence.

### Game acceptance

In the context of this research, game acceptance refers to the degree to which participants felt engaged when playing the game. We measured game acceptance by the level of enjoyment and frustration expressed after a gaming session and an overall game recommendation. At the end of each gaming session, participants were asked to use a 5-scale smiley-face Likert scale and circle their enjoyment level (1 = very boring; 5 = very fun) and perceived difficulty (1 = very easy; 5 = very hard). The means of all sessions for enjoyment and perceived difficulty were calculated as a cumulative measure of appraisal of the game participants played. At the end of the two weeks, during the debriefing session, participants were asked if they would recommend the game on a three-point scale ranging from “Yes”, “Maybe with changes” to “No”.

### Assessment of rhythmic abilities

Rhythmic abilities were assessed with selected tests from BAASTA. We selected tasks well suited to capture rhythmic abilities’ perceptual and sensorimotor dimensions (Dalla Bella et al., 2024) while being feasible for children (Puyjarinet et al., 2017; Begel et al., 2022). The tasks involved the Beat Alignment Test (BAT), paced tapping to a metronome, and paced tapping to music.

The BAT was initially designed by Iversen & Patel (2008) to assess participants’ ability to perceive the beat inherent to a musical stimulus. The participants were submitted to the abbreviated version of the BAT (24 trials, with a 600-ms inter-beat interval, instead of the full 72-stimuli version; Dalla Bella et al., 2017). Excerpts counting 20 beats each, taken from Bach’s “Badinerie” and Rossini’s “William Tell Overture”, were presented. An isochronous sequence (metronome) superimposed on the music was either aligned or not with the beat. Out of 24 trials, 16 were not aligned to the beat. The non-alignment was initiated either by a phase shift or a period shift. Participants were asked whether the sequence aligned with the music’s beat. They were given two examples of aligned and non-aligned trials, followed by four training trials. The duration of the test was approximately eight minutes.

In paced tapping to a metronome and music, participants were asked to tap their finger in synchrony with an isochronous sequence of piano tones and with short musical excerpts taken from Bach’s “Badinerie” and Rossini’s “William Tell Overture” (for details, see Dalla Bella et al., 2024; Dalla Bella et al., 2017 for the tablet version). The participants repeated the test twice for each metronome or musical stimulus. The participant was given a short training before each different tempo or excerpt. The overall duration of these tests was approximately ten minutes.

### Assessment of executive functions

We administered set-switching, Eriksen flanker, and Go/No-go tests to assess executive functioning.

### Set-Switching test

This test was organized into three different blocks (48 trials per block): “location”, “direction” and “mixed”. In the “location” block, participants had to identify where a red arrow appeared on the screen while ignoring the direction it was pointing. Conversely, in the “direction” block, participants were asked to respond only to the direction a blue arrow was pointing, regardless of which side of the screen it appeared on. The “mixed” block presented red and blue arrows in a predetermined pseudo-random order. When participants saw red arrows, they were asked to abide by the “location” rule, and when blue arrows appeared, they had to switch to the “direction” rule. Participants were invited to respond as fast as possible in all blocks. In all trials, a white fixation cross was shown for 500 ms; then the arrow was displayed until the participant responded (up to a maximum of 4500 ms), and between trials, a blank screen was shown for 750 ms. This task took approximately 6-8 minutes for each participant to complete.

### Eriksen Flanker fish task

Participants were presented with graphics of one or five fish of the same size arranged in a horizontal line on the screen. The participants were asked to respond as quickly as possible by pressing one of two keys to indicate the direction in which the middle fish was swimming (either left or right) while ignoring any other fish surrounding (flanking) the middle fish. In one-third of the trials, only one fish was presented without any flanking fish (neutral); one-third of trials contained flanking fish that were oriented in the same direction as the middle fish (congruent trials); and one-third of the trials contained flanking fish oriented in the opposite direction as the middle fish (incongruent trials). Participants had a 2000 ms (ISI) to respond, including a 500 ms stimulus and 1500 ms fixation duration. In between all trials, a white fixation cross was shown for 750 ms; then, the stimulus was shown until the participant responded. This test had 96 trials and took approximately 3-5 minutes for participants to complete.

### Go/No-Go task

In this test, participants were shown two different stimuli: an orange circle or a blue circle. The participant was asked to press the space bar on a physical keyboard as soon as possible when a blue circle appeared (Go-trial) and to refrain from pressing any key when an orange circle appeared (No-Go trial). Blue circles were shown in 70% of the trials, and orange circles in the remaining 30%. After a 1500 ms white fixation cross, the stimulus was shown on screen for 500 ms, for a total inter-onset interval of 2000 ms. Late responses (i.e., during the following fixation period) were accepted during the task, and then a response timing criterion was applied during analysis. This task used 100 trials and took approximately 4-5 minutes for each participant to complete. A minimum of 10 trials was required to be included in the analysis. Responses shorter than 100 ms were not considered valid inhibition control responses, and therefore were excluded from analysis.

### Measures and statistical analyses

All statistical analyses were performed using R Studio (R Core Team, 2022).

### Compliance and acceptance

To compare the means of both games on measures related to compliance and acceptance, we utilized the Wilcoxon rank sum test because we did not expect data for these measures to be normally distributed.

### BAASTA

We computed d-prime (*d’*) scores for the BAT task to obtain a measure of perceptual beat tracking sensitivity while accounting for response bias.

For the paced tapping tasks, logit-transformed vector length values were used to measure synchronization consistency, as described above (see “Stimulus selection”). The synchronization consistency scores for each subtask (three metronome tempos and two music excerpts) were then each converted to z-scores according to the entire sample’s pre-training mean and standard deviation for the subtask. These scores were then averaged per participant and timepoint at the task level, yielding standardized scores (i.e., derived from z-scores) of synchronization consistency for paced tapping to metronome and paced tapping to music.

Subsequently, to obtain a measure that reflects both perceptual and sensorimotor rhythmic abilities, we computed the Beat Tracking Index (BTI). The BTI was used because it has a high test-retest reliability in adults (Dalla Bella et al., 2024) and has successfully captured individual differences in children and adults with neurodevelopmental disorders (ADHD; Puyjarinet et al., 2017). The BTI was calculated by averaging z-scores of the perceptual performance (BAT d’) with the average of the scores of tapping to metronome and music.

To assess changes in rhythmic performance (post-minus pre-training) of participants in each game, we estimated linear models using the *lm()* function in R for each of the four rhythmic performance scores (BTI, paced tapping to music, paced tapping to metronome, and BAT *d’*). The dependent value for each model was the difference scores between time points, i.e., post-training performance minus pre-training on the corresponding score. Due to the inherent difference in continuous playing time (CPT) between RW and FB arising from navigation through map levels and score screens, these models included a mean-centered term for CPT to account for this variation between the games. Moreover, previous research on RW in adults with Parkinson’s disease indicated that individuals may benefit differently from playing the game based on their pre-training rhythmic abilities (Dalla Bella et al. 2018), so the models also included a term for the baseline pre-training performance score. The linear models thus contained these predictors as main effects, as well as all interactions (2-way and 3-way):

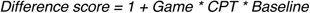

Additionally, the effect of individual differences in training dose was assessed within each game by testing the Pearson correlations between rhythm scores and training duration (total training duration and CPT, separately. Effect size interpretations of eta squared for linear regression are based on (Miles & Shevlin, 2001).

### Executive functioning tasks

Participants were excluded from analysis on any executive functioning task if one of their time points had a mean proportion of correct responses ≤ .50, representing chance or below-chance performance; this criterion excluded two participants for the Flanker task (one from each group), and one (RW) from the Set-Shifting task. A chance performance threshold may be slightly higher if calculated using permutation tests; however, given this research objective’s exploratory scope and the study’s small sample size, a .50 cut-off was considered sufficient.

For the Flanker and Set-Shifting tasks, response time (RT) values were combined into a single executive functioning index, as described below. RT values were calculated after removing trials with incorrect responses, and after removing trials with early or late responses. For both the Flanker and Set-Shifting tasks, RT values shorter than 100 ms were discarded. Additionally, for the Flanker task, RT values greater than 1300 ms were discarded, whereas for the Set-Shifting task, longer RT values were expected due to the higher complexity task and no maximum criterion was applied. For the Set-Shifting task, we then subtracted the mean RT of non-mixed blocks of the task (i.e., when only the “location” or “direction” rule was applied) from the mean RT of the mixed section (when the “location” or “direction” rule alternates pseudo-randomly). For the Flanker task, we subtracted the mean RT of congruent trials from the mean RT of the incongruent trials.

Following the approach of previous studies using the Trail-Making Test, which is a similar task involving simultaneous executive functioning processes (i.e., processing speed, inhibition, cognitive flexibility), we calculated a proportional score based on RT values. Such proportional scores are used as a sensitive index of change in executive functioning following an intervention (Periáñez et al., 2007; Stuss et al., 2001). The mean RT of the incongruent trials was subtracted from the mean RT of congruent trials, and this difference was divided by the RT of congruent trials, i.e. (Congruent RT – Incongruent RT) / (Congruent RT), where higher (less negative) scores represent higher attentional functioning. It should be noted that this formula differs from typical use of proportional scores in that the order of terms in the numerator is reversed (i.e., a typical approach would use Incongruent RT minus Congruent RT); this change allowed for higher values to represent better performance, and for higher difference scores (of proportional scores between time points) to indicate improvement after training.

We expected the experimental condition to train a combination of cognitive flexibility and inhibition control processes and, therefore, scores from the Flanker and Set-Shifting tasks were combined into a single executive functioning index. Proportional scores at pre-intervention and post-training from both tasks were converted to z-scores using the task means and standard deviations of all participants at pre-training, and then the Flanker and Set-Shifting z-scores were averaged at each time point for each participant. Improvement was calculated by computing post-minus pre-training executive functioning index scores. After graphically and statistically verifying that the scores were normally distributed between groups, we performed a one-tailed independent samples t-test to assess whether difference scores from the RW group were greater than those from the FB group. We then tested if the within-group change was significantly greater than 0 using a one-sample t-test.

On the Go/No-Go task, scoring was based on signal detection theory in order to measure the sensitivity to correctly detect “go” stimuli, while accounting for response bias or the general tendency to respond. We calculated *A’,* the non-parametric equivalent of *d’*, to avoid score approximations in the case of ceiling hit rates or zero false alarm rates, which were common in this task. An *A*’ near 1.0 indicates high discrimination sensitivity, while a value near 0.5 means chance performance (Makowski, 2018).

## Results

We begin by presenting the results of compliance and the acceptance of both games in children with ADHD, followed by the effects of RW training on rhythmic and executive functioning.

### Compliance and acceptance

A total of 87.1% (27 out of 31) of the children with ADHD completed the study protocol. Four children, two from the RW group and two from the FB group, did not complete the study due to time management issues or severe ADHD symptom manifestations, and are not included in analyses. Additionally, one participant was excluded from all analyses due to a technical problem where the participant accidentally changed the tablet’s settings, affecting the game’s progression and invalidating their training progress. One participant had unreliable logged data regarding training compliance, so their self-reported training duration was used instead of their logged training duration. All remaining participants completed seven to ten (the maximum) self-reported questionnaires about each gaming session and returned the tablet data.

As shown in Table 3, participants in both groups showed similar total training duration progress (88% for RW and 100% for FB), meeting the target set for 300 minutes of training over two weeks. Despite these similarities, we found that the amount of CPT in the RW group (M = 141 minutes, SD = 60, Range = 30-244) was significantly lower than the amount of CPT in the FB group (M = 275, SD = 83, Range = 103-367; W_A_ = 19, p < 0.001). The CPT was 54% of the logged training duration in RW and 86% in FB. Training duration was strongly and positively correlated with the CPT, r(24) = .73, p < .0001, suggesting both variables reliably measure training dose.

**Table 3.** Compliance with protocol targets and acceptance for the two serious games.

| Characteristic | Rhythm Workers |  |  |  |  | Frozen Bubble |  |  |  |  | p-value |
| --- | --- | --- | --- | --- | --- | --- | --- | --- | --- | --- | --- |
|  | N | Mean | SD | Min | Max | N | Mean | SD | Min | Max |  |
| Training Minutes | 13 | 267.0 | 96.3 | 87.5 | 417.7 | 13 | 321.8 | 94.4 | 135.9 | 454.9 | 0.2 |
| Taps per Minute<sup>1</sup> | 13 | 61.2 | 9.1 | 44.9 | 78.8 | 13 | 56.6 | 12.5 | 39.3 | 83.9 | 0.3 |
| Game Enjoyment | 12 | 3.5 | 0.6 | 2.3 | 4.5 | 13 | 3.5 | 1.0 | 1.6 | 5.0 | 0.9 |
| Perceived Diff. | 13 | 3.7 | 0.7 | 1.9 | 4.8 | 12 | 2.8 | 0.8 | 1.6 | 4.2 | **0.03\*** |
| Recommend | 13 |  |  |  |  | 13 |  |  |  |  | 0.9 |
| “Yes” |  |  | n = 6 (55%) |  |  |  |  | n = 6 (75%) |  |  |  |
| “Maybe with changes” |  |  | n = 2 (27%) |  |  |  |  | n = 1 (13%) |  |  |  |
| “No” |  |  | n = 2 (18%) |  |  |  |  | n = 1 (13%) |  |  |  |
| Did not answer |  |  | n = 3 |  |  |  |  | n = 5 |  |  |  |

<sup>1</sup>Total number of logged finger-taps by game divided by training duration in minutes

Comparable amounts of motor movement (number of taps) were observed between games as shown by similar tapping rates per minute of training between games (p = .29). Participants expressed above-average levels of enjoyment, comparable for both games (p = .98). When participants were asked if they would recommend the game, more than half of the participants in each group responded positively, and the distribution of responses was similar between games (p = .90). Participants in the RW group perceived the game as significantly more difficult than FB (W_A_ = 119, p = .03); however, perceived difficulty did not correlate significantly with enjoyment in RW (p = .80) or FB (p = .09).

Both groups played a comparable number of sessions, averaging 5 per week per game. Figure 3 shows the evolution of enjoyment and perceived difficulty over the sessions for both games.

**Figure 3.**
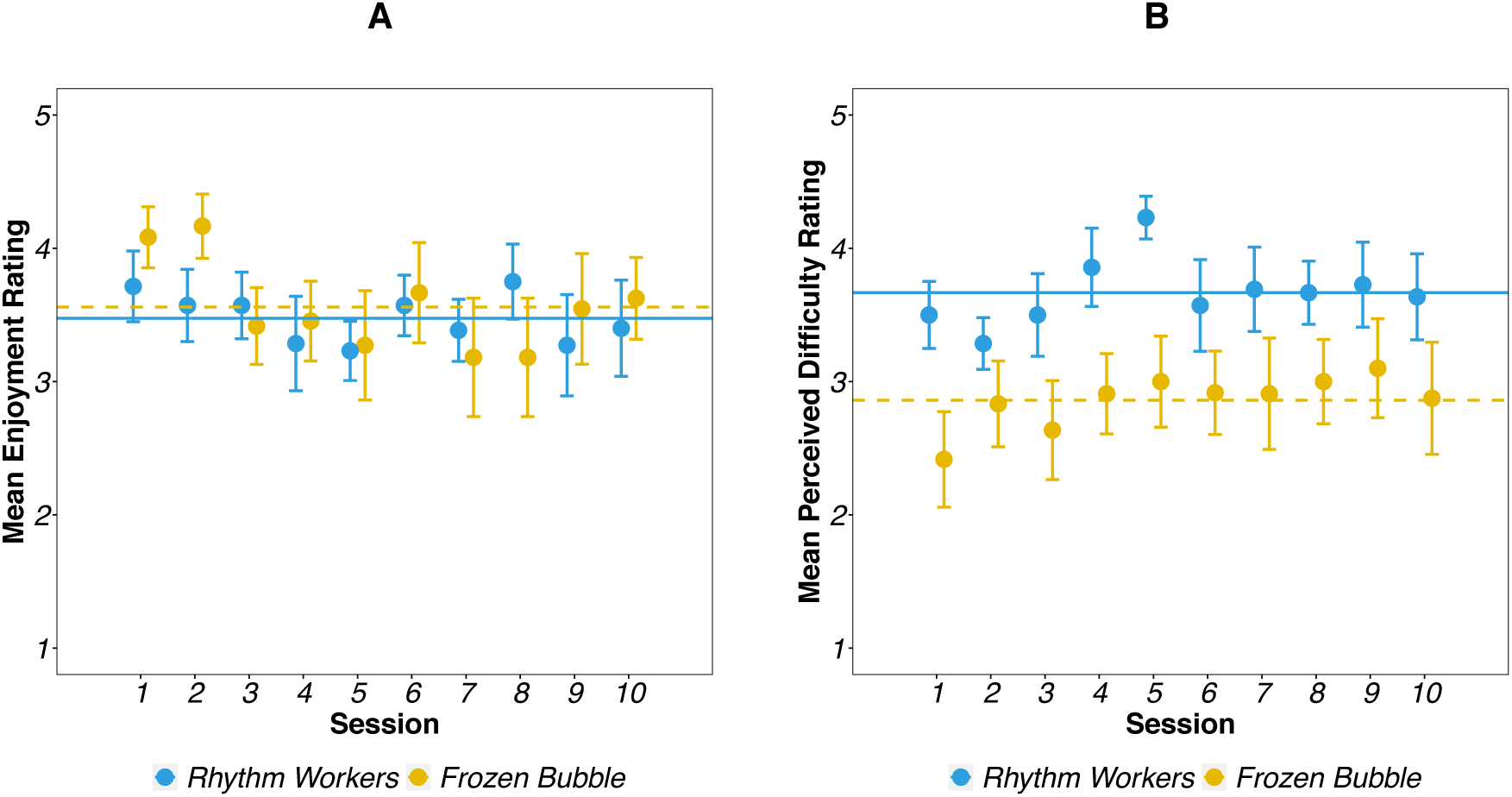
Time series of mean enjoyment (A) and mean perceived difficulty over the sessions (B) between Rhythm Workers and Frozen Bubble. Error bars indicate the standard error, and horizontal lines represent the means across sessions.

### Rhythmic performance

Table 4 shows the rhythmic performance at both time points between games on individual tasks of BAASTA and the BTI.

**Table 4.** Rhythmic Performance for children with ADHD after training.

| Rhythmic Domain |  | Rhythm Workers |  |  |  |  | Frozen Bubble |  |  |  |  |
| --- | --- | --- | --- | --- | --- | --- | --- | --- | --- | --- | --- |
|  |  | N<sup>2</sup> | Mean | SD | Min | Max | N<sup>2</sup> | Mean | SD | Min | Max |
| BTI | *Pre* | 13 | 0.06 | 1.04 | -1.25 | 1.90 | 12 | 0.14 | 0.70 | -0.84 | 1.31 |
|  | *Post* | 13 | 0.21 | 0.91 | -1.43 | 1.51 | 12 | 0.10 | 0.71 | -0.68 | 1.22 |
| Paced Tapping to Music<sup>1</sup> | *Pre* | 12 | 0.13 | 0.94 | -1.12 | 1.58 | 11 | -0.20 | 0.98 | -1.71 | 1.18 |
|  | *Post* | 13 | 0.23 | 0.82 | -1.12 | 1.33 | 12 | -0.41 | 1.16 | -2.19 | 1.08 |
| Paced Tapping to Metronome<sup>1</sup> | *Pre* | 13 | 0.41 | 0.90 | -0.77 | 2.34 | 12 | 0.44 | 0.80 | -0.93 | 1.59 |
|  | *Post* | 13 | 0.42 | 1.25 | -3.35 | 1.90 | 12 | 0.45 | 0.88 | -0.88 | 1.83 |
| Rhythm Perception *d'* | *Pre* | 13 | 1.24 | 1.19 | -0.29 | 3.54 | 12 | 1.70 | 0.8 | 0.74 | 3.07 |
|  | *Post* | 12 | 1.55 | 1.09 | -0.16 | 3.07 | 12 | 1.74 | 0.76 | 0.76 | 2.79 |

<sup>1</sup> Synchronization consistency score
<sup>2</sup> Due to missing data for some participants who could not complete all the tasks because of task difficulty (e.g., tapping in antiphase or double-time) or task-specific inattention

Figure 4 shows the difference scores collected for the BTI and each component of the BTI.

**Figure 4.**
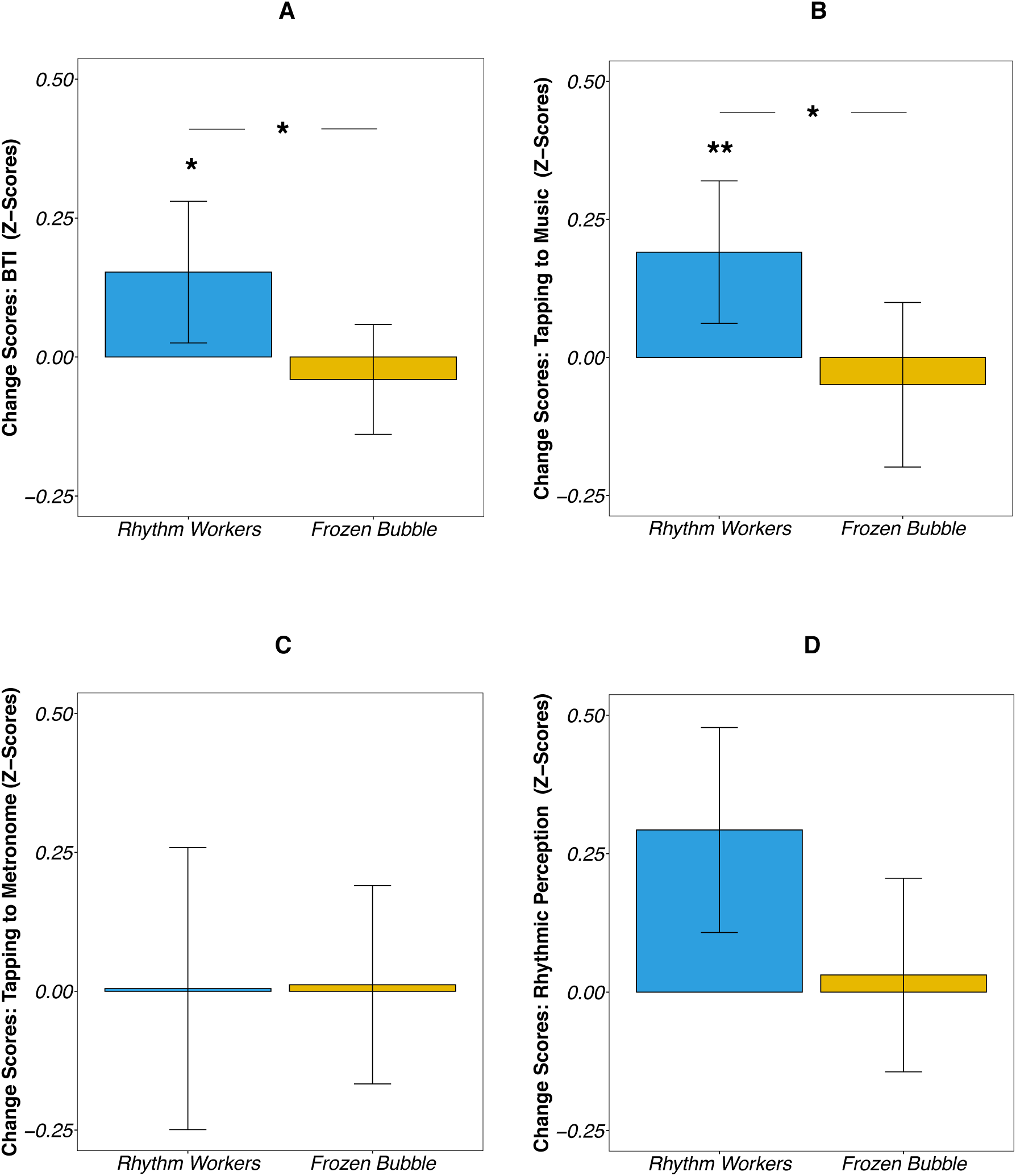
Difference scores were obtained for the BTI (A) and each of its components: paced tapping to music (B), paced tapping to a metronome (C), and rhythm perception (D). Bars indicate means and the standard error; * p <.05; ** p = < .01.

In the BTI, which captures the general capacity to track a beat, a positive effect of the training condition (t(17) = 2.34, p = .031) was found for those who played the RW game (difference score, M = .15, SD = 0.46) compared to those who played the FB game (difference score, M = -.04, SD = .34). We also observed within-group improvement for RW, as indicated by a significant model intercept (t(17) = 2.87, p = .01). No interactions terms in the model were significant (p ≥ .27) and the omnibus model provided an overall good fit (F(7, 17) = 2.056, p = .11, R^2^ = .46), with a medium effect size (η² = 0.06).

Similarly, for tapping to music there was a positive effect of the training condition (t(15) = 2.80, p = .015) for those who played the RW game (difference score, M = .19, SD = 0.47) compared to those who played the FB game (difference score, M = -.05, SD = .52). We also observed within-group improvement for RW, as indicated by a significant model intercept (t(15) = 3.04, p = .008). No interaction terms in the model were significant (p ≥ .19), and the omnibus model provided an overall good fit (F(7, 15) = 2.69, p = .051, R^2^ = .56), with a medium effect size (η² = 0.06).

The omnibus models were not statistically significant for paced tapping to a metronome (p = .46) and the BAT rhythm perception task (p = .62). Rhythm perception improvements were not found to be correlated with improvements in beat-synchronization tasks in this sample (p = .56).

To assess whether the positive effect of the rhythmic training was linked to the individual amount of training, we tested the relation between CPT and rhythmic improvements (see Figure 5). For RW, CPT was strongly and positively correlated with improvement in paced tapping to music (r(10) = .68, p = .008) and the BTI (r(10) = .50, p = 0.04), but not for FB (p ≥ .16). We found a similar trend for total training duration, which was positively correlated with improvements in paced tapping to music (r(10) = .63, p = .01) and marginally significant for the BTI (r(10) = .45, p = .06) for RW group, but not FB (p ≥ .14).

**Figure 5.**
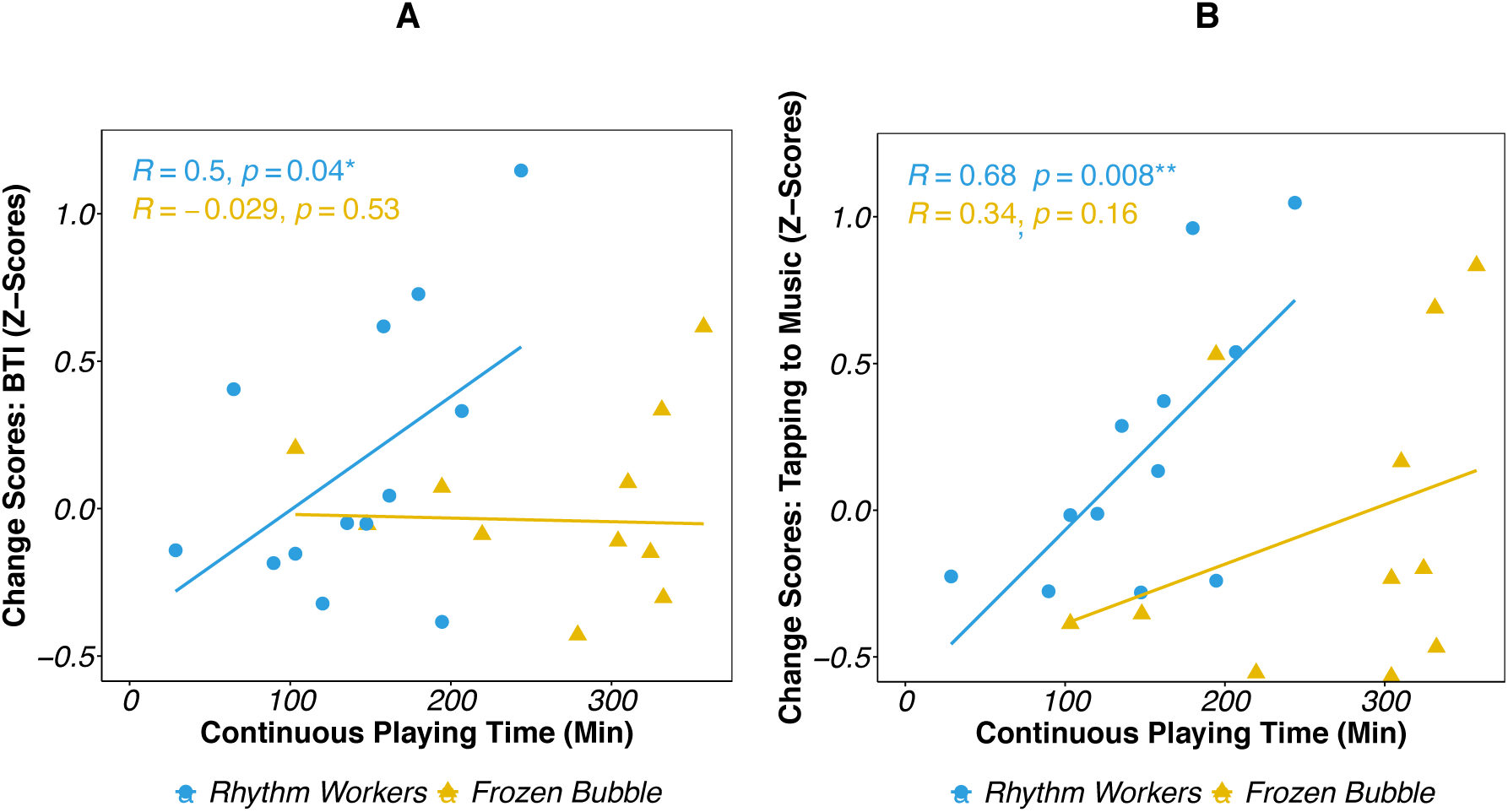
Relationship between training dose and improvement. In (A) the Beat Tracking Index and (B) the paced tapping to music task by game.

### Executive functioning

Figure 6 presents the difference scores between each game for the executive functioning index.

**Figure 6.**
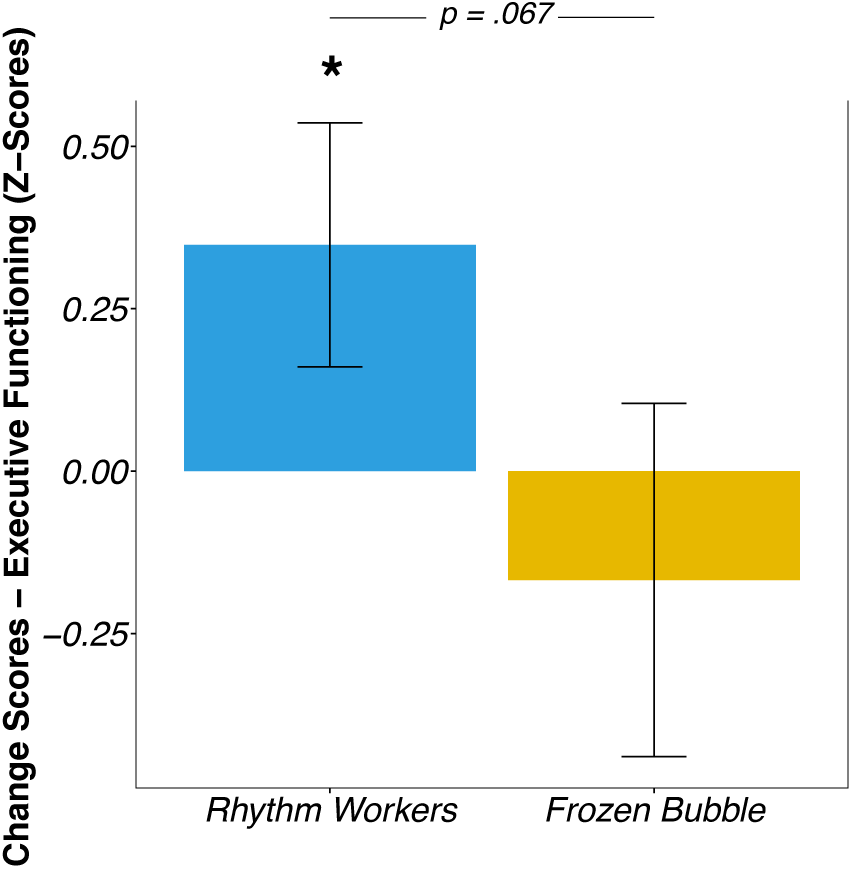
Difference scores obtained for the executive functioning index - Bars indicate means and standard errors; * p <.05

Training with RW had a positive effect on proportional RT scores of the combined executive functioning index (difference score, M = .35, SD = .65) compared to playing FB (difference score, M = −0.17, SD = .98); the difference between training conditions just failed to reach statistical significance (t(22) = 1.56, p = .067, Cohen’s *d* = .63). Children who played RW showed an improvement which was significantly greater than zero (t(11) = 1.86, p = .045); this was not the case for FB (p = .73). There was no difference between games on the A’ difference scores of the Go/No-Go (p ≥ .65).

## Discussion

We conducted a proof-of-concept study to test gamified rhythmic training for children with ADHD employing a parallel-arm randomized control trial design. The protocol was carried out remotely using digital training, online tools, and equipment mailing. The rhythmic “serious game” on a tablet, RW, was compared to an active control condition (non-rhythmic serious game, FB). The primary goal was to assess the feasibility of the training program in children with ADHD, whose impairments in attention are linked to rhythmic deficits (Puyjarinet et al., 2017). A secondary goal was to test the effectiveness of the training on rhythmic abilities using BAASTA and, as an exploratory aim, on executive functioning.

Participants achieved an 87% completion rate of the entire research protocol, surpassing the expected rate (70%) for pediatric mental disorders (Bogdan et al., 2023). The target training duration of 300 minutes was met at 88% for RW and 100% for FB. Both games exhibited similar enjoyment levels and comparable amounts of motor activity (finger taps). Enjoyment values were high and around 3.5/5. Therefore, we expect that children would be willing to play the game in a more extended training program while maintaining their enjoyment. Despite RW being perceived as more difficult, this was not related to the enjoyment children experienced during the 2-week training sessions; moreover, this did not impact the improvement of sensorimotor or executive functioning. It is noteworthy that participants spent more continuous time playing FB due to RW’s level navigation time. Each level in RW ranged between 1-3 minutes; then, participants spent time on the score, medal, and map screens between levels. This imbalance worked in favor of the study’s research objectives and showed that even less training than anticipated is sufficient for producing specific training effects. Another encouraging statistic was that only 18% of responding participants who played RW and 13% of those who played FB did not recommend the game they played. This proof-of-concept assessment provides a suitable empirical basis for analyzing and interpreting the training’s effectiveness in improving rhythmic and executive functioning abilities. The control game FB is particularly well suited for the current research objectives and an excellent comparison to draw precise causal inferences related to training-induced improvements in other domains.

### Effects of a “serious” rhythmic game on rhythmic abilities in ADHD

This study presents the first empirical evidence that gamified rhythmic training can selectively enhance rhythmic abilities in children with ADHD, a population characterized by rhythmic impairments (Noreika et al., 2013; Puyjarinet et al., 2017). The effect of the rhythmic training was observed in improved paced tapping to music and rhythmic perception in BAASTA tasks (Dalla Bella et al., 2024). No comparable improvement was observed with the non-rhythmic video game (FB). Notably, time spent on RW correlated strongly with auditory-motor improvements, extending to beat-tracking abilities (Beat Tracking Index – BTI; Puyjarinet et al., 2017) involving a broader set of sensorimotor skills. This study marks the first use of BTI in neurodevelopmental disorders with a randomized-control trial design, validating BAASTA for detecting sensorimotor training effects in ADHD within two weeks. Consistent with previous findings in persons with Parkinson’s disease (Dalla Bella et al. 2018), individuals with stronger initial rhythmic skills benefited more from the training, suggesting a potential parameter for adaptive game design in future interventions.

### Effects of a “serious” rhythmic game on executive functions in ADHD: preliminary evidence

Beyond observing sensorimotor improvement, we investigated the transfer of skills from sensorimotor synchronization to executive functioning. Despite the limited training time compared to full-fledged music training programs (300min-19500min; Jamey et al., 2023), the current findings show preliminary evidence for improved executive functioning in children trained with RW, but not in those who played FB. Compared to FB, the executive functioning change difference after RW training was substantial, with a medium effect size (d = .63), and 93.3% likely not due to chance. This executive functioning improvement, comprising inhibition control and cognitive flexibility tasks, showed speed but not accuracy improvements, which were above 90% in both groups. The Go/No-Go task showed no game-specific improvement due to ceiling performance before the training. These preliminary results are promising and might lead to even more prominent (and statistically significant) effects of rhythmic training on EF with a longer training duration and a larger sample size. In sum, this study offers early causal evidence that rhythmic training benefits may transfer across different cognitive and motor abilities in children with ADHD and pave the way to innovative training applications.

The predictive timing of associated cognitive and neural processes can explain the potential transfer of rhythmic training effects across motor and executive function systems. Predictive timing involves accurately anticipating when a future event will occur based on consistent temporal patterns. Researchers like Large & Jones, 1999; Piras & Coull, 2011; Schwartze & Kotz, 2013) have extensively discussed this concept and its underlying mechanisms, which include connections between subcortical and cortical brain regions. Rhythmic training, such as gait rehabilitation with auditory cueing, is likely to activate these neural networks. Explicit timing tasks are considered a form of “cognitively controlled timing,” and rely on basal-ganglia-cortical networks. This auditory rhythm-based training may capitalize on the preserved predictive timing observed in individuals with ADHD, potentially improving with training. This aligns with findings that rhythmic training enhances beat tracking, positively affecting executive functions. Notably, rhythmic training’s benefits appear linked to a central predictive mechanism involving executive functioning, as indicated by Puyjarinet et al.’s (2017) study correlating timing deficits in ADHD with impaired executive functioning.

### Limitations and future directions

This proof-of-concept study has methodological and statistical limitations. Technical issues affected one participant’s game performance, highlighting the need for future prototypes to hardwire device latency for specific tablet models. The sample size was tailored for proof-of-concept, limiting the scope of analyses. A larger cohort is recommended for extrapolating and replicating findings, enabling more sophisticated statistical analyses. Modification of the Go/No-Go task’s difficulty is suggested, as it reached ceiling performance. The study lacked double-blinding verification, but the methodology is innovative for music training because it can technically afford a certain degree of double-blinding. When more general music training is administered, it can be difficult for a laboratory specialized in music and psychology to hide the research objective from participants. By including music in both conditions, the more subtle manipulation of synchronization demands conceals the research objective more effectively.

Additionally, we did not measure the ADHD symptom levels or types of our participants beyond the presence of an ADHD diagnosis. To better understand how individual differences in symptom severity may affect the effectiveness of the training, we recommend future studies to incorporate a measure of clinical severity. However, the current study sets the stage for a randomized clinical trial with a larger sample. Future research will explore structural and functional brain changes, shedding light on perceptual and motor processes in rhythmic performance. Longitudinal studies with larger cohorts will examine the potential long-term effects of rhythmic training using “serious games” on social activities such as conversational skills and family quality of life.

This rhythmic training format could benefit other clinical populations, such as children on the autism spectrum (AS) who exhibit poor predictive timing, cerebellar dysfunction, and executive function impairments linked to social and communication challenges (Kelly et al., 2021). Children on the AS often face difficulties adhering to therapy schedules (Sandoval-Norton & Shkedy, 2019), and the minimally social rhythmic “serious game” format may be particularly engaging for them. Extending similar rhythmic training to children on the AS could explore whether improvements in executive functioning mediate enhancements from sensorimotor skills to core symptoms like speech and communication deficits. Another relevant group is children who stutter, as evidence suggests non-verbal sensorimotor timing deficits in adolescents with higher levels of stuttering severity (Falk et al., 2015). Applying the protocol to stuttering children may yield beneficial outcomes for restoring specific speech functions.

## Conclusions

RW and the control game FB were successfully validated in children with ADHD. The current results provide initial causal evidence that playing RW improves these children’s sensorimotor synchronization and specific executive functions (EFs). Training duration of approximately 300 minutes was sufficient for observing these improvements, with a positive correlation between training duration and the strength of improvement in sensorimotor synchronization. These findings suggest that mechanism-driven approaches in music training are efficient for enhancing non-musical skills, requiring a lower dose to generate effects, and enabling tailored training for specific clinical populations. Future investigations exploring underlying neural mechanisms and increased statistical power are encouraged to validate these findings further and assess skill transfer effects in neurodevelopmental disorders.

## Data Availability

All data produced in the present study are available upon reasonable request to the authors.

## Declarations

## Funding

This work was supported by funding from grant 05453 from the Natural Sciences and Engineering Research Council of Canada (NSERC), grant 115050 from the Canadian Institute of Health Research (CIHR), and grant 0160 from NSERC Tier 1 Canada Research Chairs to Simone Dalla Bella, Canada Research Chair in Music Auditory-Motor Skill Learning and New Technologies.

## Conflicts of interest/Competing interests

SDB is on the board of the BeatHealth company dedicated to the design and commercialization of technological tools for assessing rhythm abilities such as BAASTA tablet and implementing rhythm-based interventions. Other authors have no competing interests to disclose.

## Ethics approval

Ethical approval for this research was granted by the Comité d’éthique de la recherche en éducation et psychologie (CEREP) of the Université de Montréal - CEREP-20-008-P.

## Consent to participate

All participants consented to participate.

## Consent for publication

All co-authors consent to publication.

## Availability of data and materials

The datasets generated during and/or analysed during the current study are not publicly available due to participant confidentiality but are available from the corresponding author on reasonable request. Because of this limitation, analysis code is illustrated with a synthetic dataset, which allows readers to check the correctness of their implementation. See open practices statement for links to stimuli.

## Code availability

Upon request to authors.

## Open Practices Statement

Stimuli and gameplay examples are available at: https://osf.io/gykjd/?view_only=26be79399f1e48fe838107df437e0ceb

The data and other materials for this proof-of-concept may be made available upon request to the authors. This proof-of-concept study was not preregistered.

## Notes

### Author Declarations

Ethical approval for this research was granted by the ComitE d'Ethique de la Recherche en Education et Psychologie (CEREP) of the University of Montreal - CEREP-20-008-P.

